# Within- and Between-Family Validation of Nine Polygenic Risk Scores Developed in 1.5 Million Individuals: Implications for IVF, Embryo Selection, and Reduction in Lifetime Disease Risk

**DOI:** 10.1101/2025.10.24.25338613

**Authors:** S Cordogan, DB Starr, NR Treff, J Lanchbury, E Goldstein, K Sadeghi, M Burmeister, L Dayani, P Macdonald, JD Keen-Kim, S Fishel, E Cervantes, L Folkersen

## Abstract

Polygenic risk scores (PRSs) can reduce lifetime disease risk by guiding embryo selection during in vitro fertilization (IVF). We performed genome-wide association meta-analyses totaling ∼1.5 million individuals to construct state-of-the-art PRSs for nine diseases: Alzheimer’s disease, breast cancer, coronary artery disease, endometriosis, hypertension, prostate cancer, rheumatoid arthritis, type 1 diabetes, and type 2 diabetes. The resulting predictors achieved liability-scale R^2^ values of up to 22.9% for type 2 diabetes, matching or exceeding previously published benchmarks across all scores. Three PRS - Alzheimer’s disease, prostate cancer, and type 2 diabetes - explained over 75% of the common-SNP heritability. Within-family validation in 40,872 siblings across 18,840 families showed that, for eight of nine diseases, predictive performance was comparable to population-level results, confirming substantial direct genetic effects. Modeling of embryo selection suggests that couples with five euploid embryos could achieve 27-67% relative risk reduction across diseases. While the limitations of genetic data availability meant that these estimates were performed in European ancestry samples, we performed validation in the multi-ancestry US-based All of Us Biobank, demonstrating significant statistical power across ancestries. These findings support the clinical applicability of PRS-guided embryo selection to reduce the burden of common diseases.

## Introduction

Embryos from in vitro fertilization (IVF) cycles have historically been ranked for transfer using morphological criteria and chromosome copy number screening. While morphology, and to a greater degree chromosomal euploidy, are associated with live birth rate (1), they give no window into the disease risks of the ultimate individual. A large proportion of individual disease risk can be attributed to common genetic variance, and predicted using polygenic risk scores (PRSs) (2). By applying PRSs to embryonic genetic data, parents undergoing IVF can rank and select embryos with relatively lower risk for common disease (3,4,5).

Polygenic risk scores (PRSs) are statistical calculations that estimate an individual’s genetic predisposition to developing complex diseases, such as heart disease, cancer, and mental health conditions, made possible by Genome-Wide Association Studies (GWAS). GWAS test millions of common genetic variants for association with diseases or trait status, like prostate cancer or height in large population sets. The requisite data for GWAS is found in large biobanks, such as the UK Biobank (UKBB) (6), Finngen Biobank (7), Million Veterans Program Biobank (MVP) (8), and AllofUs Biobank (AoU) (9). Each of these biobanks contain the genotype data for hundreds of thousands of people, alongside their medical records, survey responses, laboratory test results, and physical measurements.

Although GWAS has detected many genetic variants with statistically significant associations to diseases or traits, the majority of these variants have modest effect sizes and thus have limited ability to predict diseases or traits individually. However, many associated risk variants can be aggregated together into PRSs, which can be used to explain larger proportions of disease risk. PRS research and development has made significant advances improving clinical applicability (10,11) and test performance over time (12-15).

PRSs can be used to predict phenotypes from germline DNA in both adults and human embryos. On average, couples undergoing IVF generate 3 - 5 euploid embryos that are genetically viable for implantation and are available for genetic testing for disease risk (16). As such, PRSs can be used to identify the embryo with the lowest genetic risk for common, chronic diseases.

Here, we present best-in-class PRSs for nine common human diseases, constructed using GWAS summary statistics derived from sample sizes approaching 1.5 million individuals across diverse cohorts. We evaluate the predictive accuracy and cross-ancestry performance of these models and simulate their application to embryo selection scenarios. Specifically, we demonstrate that selection among sibling embryos using these PRSs can yield relative reductions in predicted genetic disease risk that are comparable to other public health initiatives.

## Results

We constructed nine disease PRSs using GWAS summary statistics generated by imputing and meta-analyzing individual GWAS studies from Million Veterans Program (MVP), Finngen, the UK Biobank (UKBB), All of Us (AoU), and various trait-specific consortia (Methods).

Performance was evaluated using hold-out samples of European-ancestry UKBB and AoU individuals, as well as AoU individuals of majority African, Admixed-American, East Asian, and South Asian ancestry. Predictive performance was evaluated as incremental Liability R^2^, also known as liability-scale variance explained, calculated using methods from Lee *et al.* (17), and lifetime prevalences derived from literature (Supplementary Table 2). Higher Liability R^2^ corresponds to increased statistical power to predict disease. The performance of rheumatoid arthritis and type 1 diabetes PRSs are likely underestimated due to the use of population prevalence, as lifetime prevalence data was not available. Disease population prevalence is generally lower than lifetime prevalence due to reduced life expectancy and increasing disease prevalence with age.

Below we compare the performance of our nine PRS to overlapping PRS from two research groups, Thompson *et al*. and Mars *et al*. (18,19). We observed improved performance across all models.

**Figure 1.**
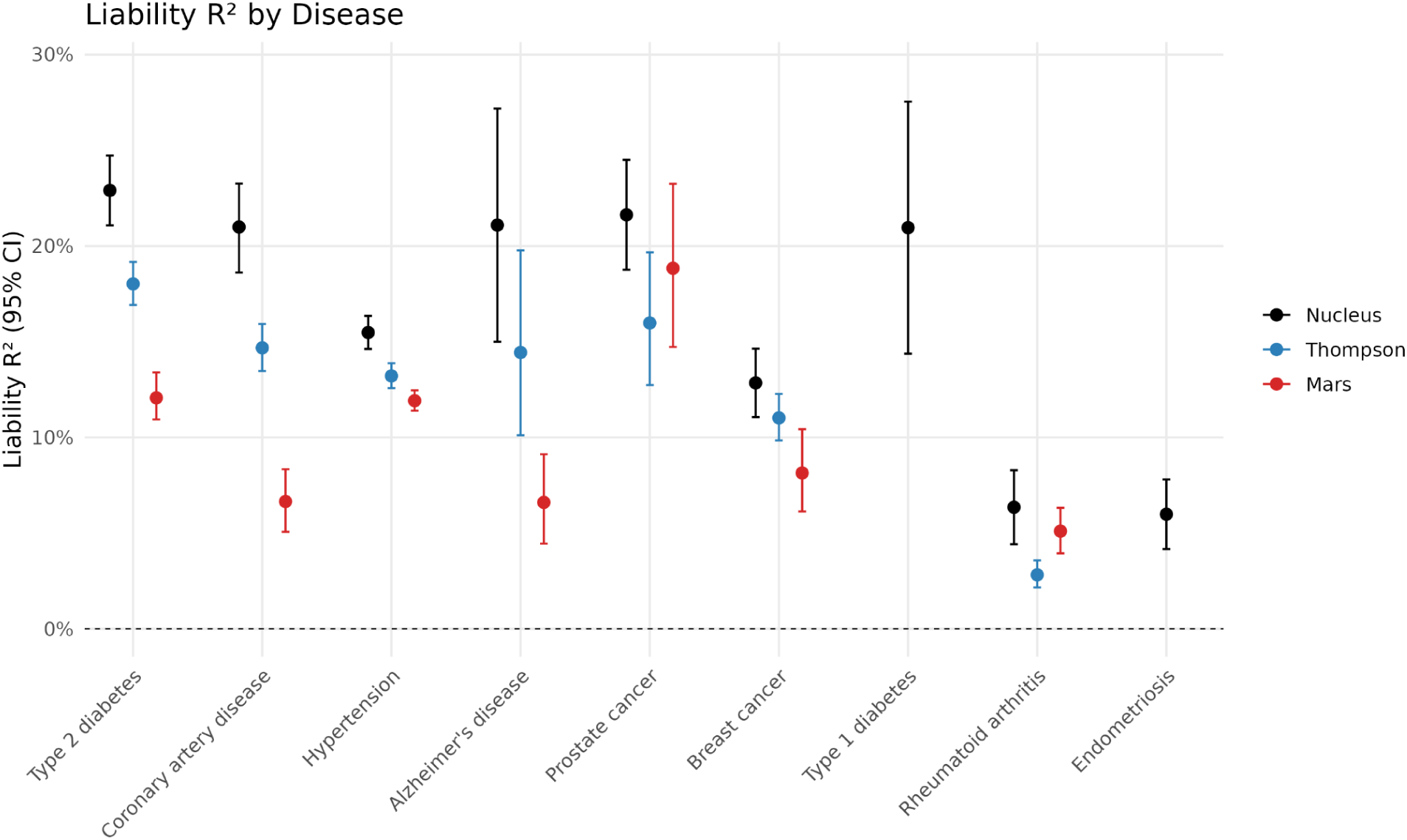
Liability R^2^ of nine PRSs, calculated on our UKBB test cohort using methods from Lee *et al*. Liability R^2^ of scores from Mars *et al.* and Thompson *et al*. were included for comparison (18,19). Type 1 diabetes was removed from Thompson *et al*. due to incompatible phenotype definitions.

The Liability R^2^ of scores was also assessed across four superpopulations within AoU for phenotypes with over 150 cases. The decreased performance of polygenic risk scores derived from majority-European GWAS within non-European populations is a well-known limitation of PRS models (20).

**Figure 2.**
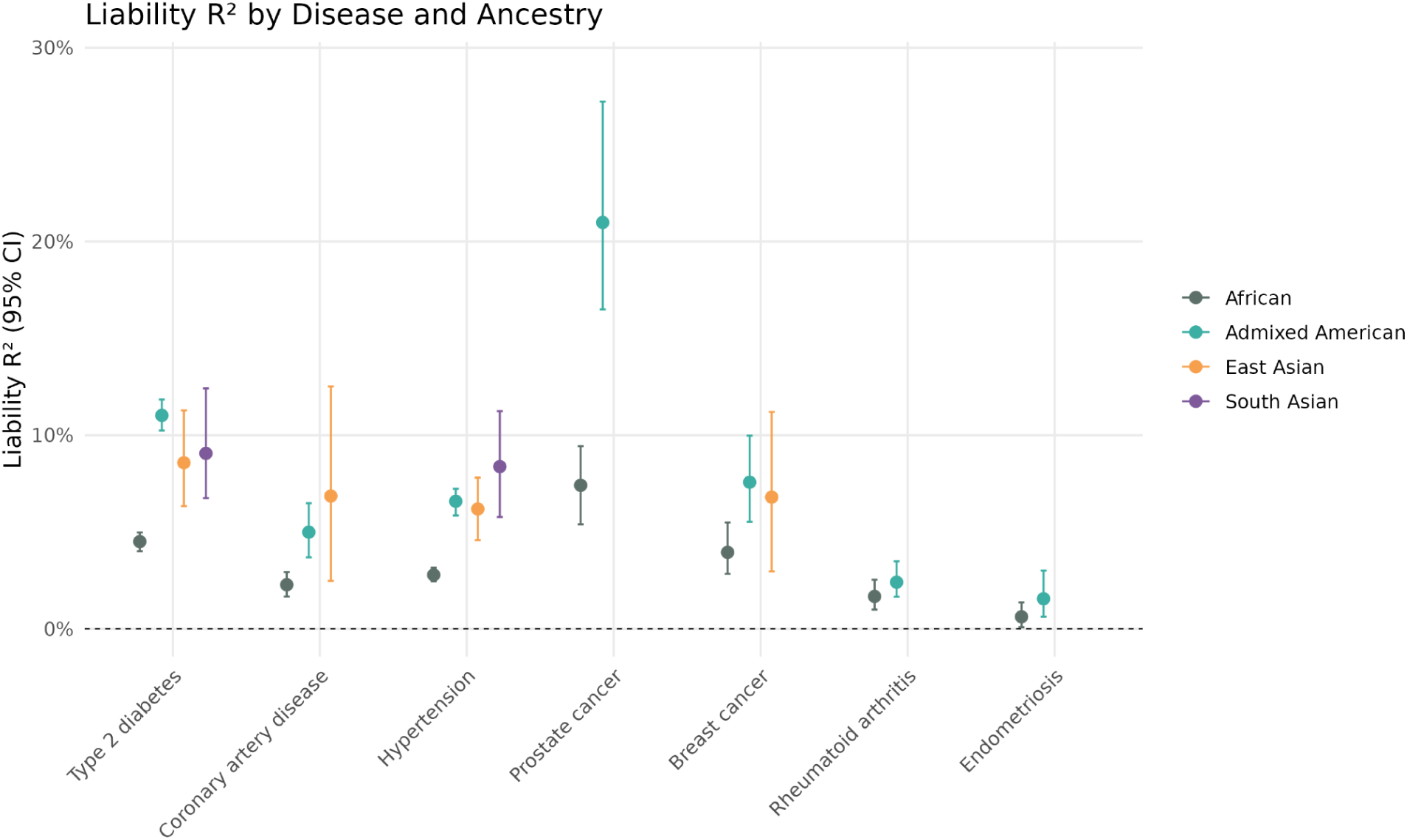
Liability R^2^ of nine PRSs within four AoU subpopulations.

Associations between genetic variants and diseases can be biased by factors beyond their direct effects on underlying individual biology, commonly known as indirect effects (21). Indirect effects include factors such as shared family environment (where parent’s genes influence children’s environment through parenting style or socioeconomic factors), population stratification (systemic differences in phenotypes between population groups), and assortative mating (when individuals mate with more genetically similar individuals), and these biases can confound PRSs. To isolate the direct genetic effects of the PRS, validation can be performed within sibling cohorts. Within these cohorts, shared family environment, population stratification, and the effects of assortative mating are held constant across siblings, allowing for a more accurate assessment of the PRS direct genetic effects.

Within-family validation was performed in a sibling cohort of 40,872 individuals across 18,840 families within the UKBB. We adapted methods for within-family PRS validation from Selzam *et al*. for binary phenotypes (21), and calculated the ratio of within-family to population performance. We found that the within-family predictive power of the PRSs was not significantly different from the population predictive power for eight out of nine scores, with the exception of type 1 diabetes. The Liability R^2^ of type 1 diabetes was reduced accordingly for risk reduction calculations in the context of embryonic selection.

**Figure 3.**
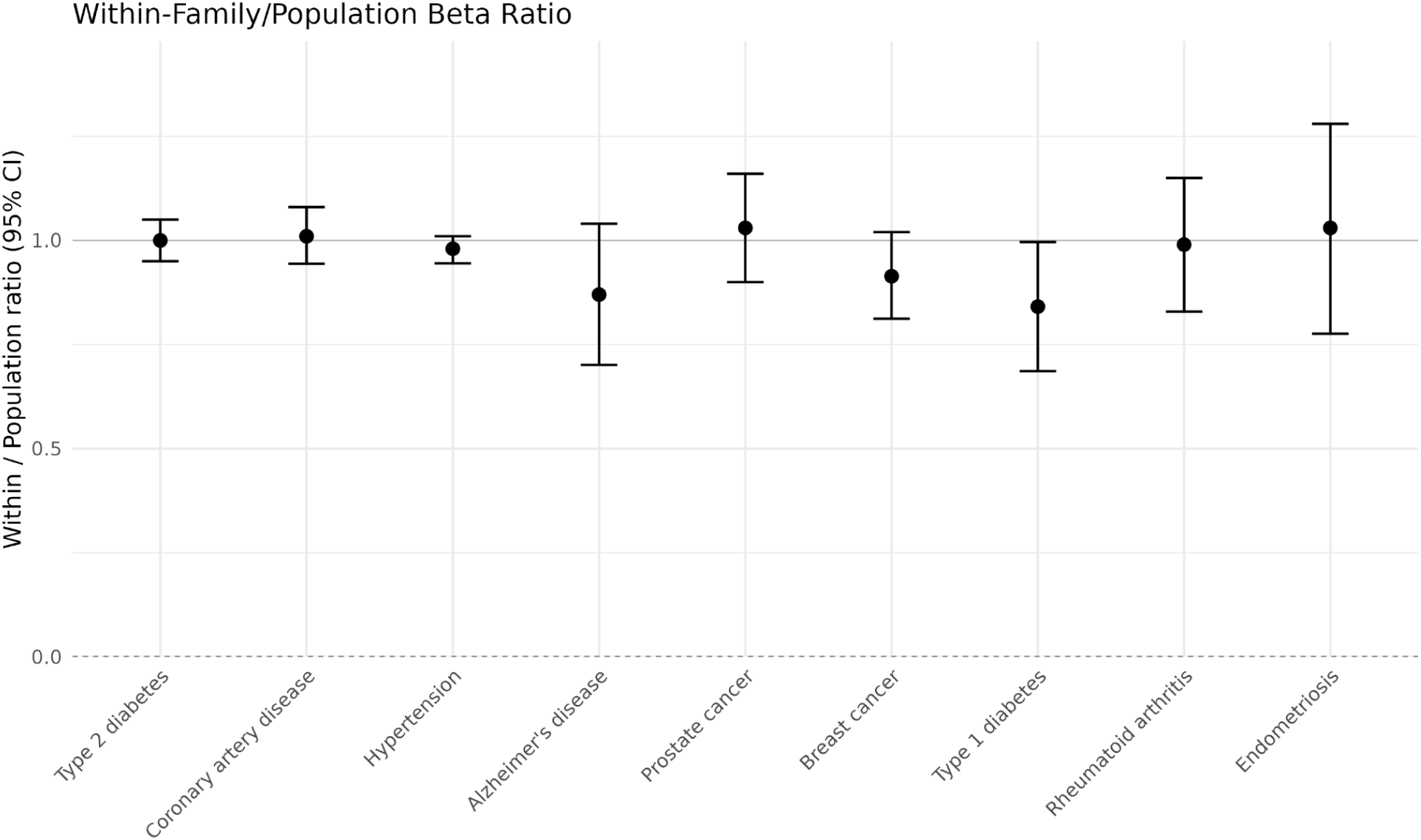
Within-family/population beta ratios of nine PRSs. For all phenotypes except type 1 diabetes, the relative performance of scores within-family was not statistically significantly different than within-population.

Further, we calculate the expected relative risk reduction from selecting a sibling embryo with the lowest disease PRS relative to the average sibling embryo, from two, five, and ten sibling embryos, using methods developed in Lencz *et al*. (5). Our PRSs achieve risk reductions greater than 50% for several diseases within a set of five siblings.

### Relative Risk Reductions within Two, Five and Ten Embryos

**Table 4.** Relative risk reductions for nine PRSs within sets of two, five, and ten sibling embryos. Relative risk reductions were calculated using lifetime prevalence for all diseases excluding rheumatoid arthritis and type 1 diabetes, where population prevalence was used as lifetime prevalence data was not available.

|  | Two Embryos | Five Embryos | Ten Embryos |
| --- | --- | --- | --- |
| Alzheimer's Disease | 31% | 55% | 67% |
| Breast Cancer | 23% | 42% | 53% |
| Coronary Artery Disease | 18% | 35% | 45% |
| Endometriosis | 17% | 31% | 40% |
| Hypertension | 13% | 27% | 36% |
| Prostate Cancer | 30% | 54% | 66% |
| Rheumatoid Arthritis | 23% | 42% | 52% |
| Type 1 Diabetes | 41% | 67% | 77% |
| Type 2 Diabetes | 27% | 50% | 61% |

Finally, we compared the Liability R^2^ to the common-SNP heritability. Common-SNP heritability refers to the total variance in disease risk explained by common genetic variants; effectively the upper bound of a PRS (2). Our three most predictive models–those for Alzheimer’s disease, prostate cancer, and type 2 diabetes–capture approximately 75% or more of the corresponding common-SNP heritability estimates. When calibrated to our lifetime prevalence assumptions, the total common-SNP heritabilities for these diseases are estimated at 24%, 28%, and 29%, respectively, compared to the 21.1%, 21.6%, and 22.9% liability-scale variance explained by our PRSs (22,23,24).

**Figure 4.**
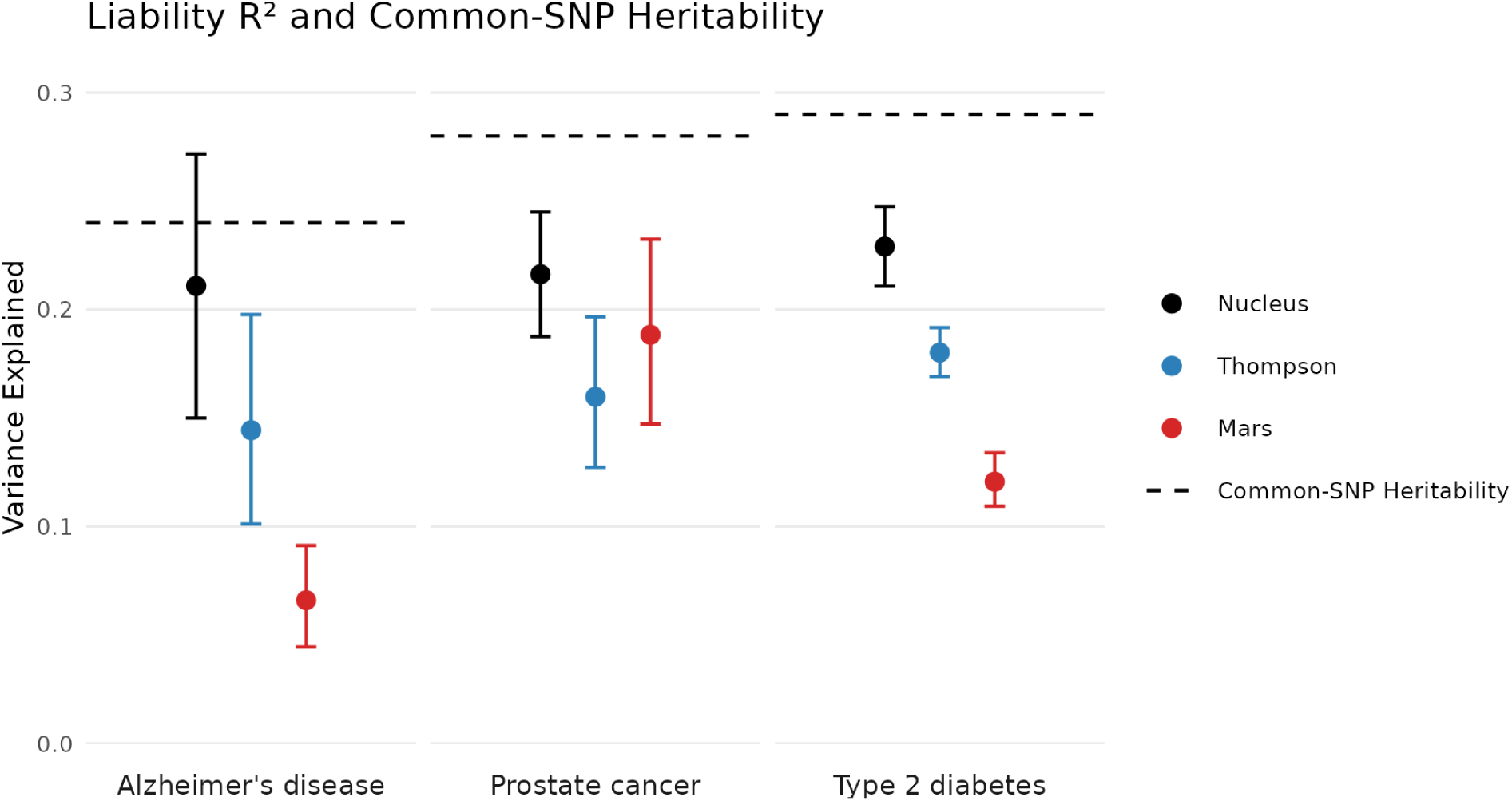
Liability R^2^ of nine PRSs, along with corresponding Thompson, and Mars PRS plotted against Common-SNP Heritability estimates for three diseases.

## Discussion

We developed and validated state-of-the-art polygenic risk scores (PRSs) for nine common diseases using genome-wide association (GWAS) data encompassing nearly 1.5 million individuals. These models were rigorously evaluated across five independent cohorts, collectively spanning five major ancestry groups and hundreds of thousands of individuals. By achieving high liability-scale predictive performance and demonstrating robust within-family validity, our findings establish that PRSs can meaningfully capture heritable disease liability even when evaluated among genetically related individuals.

Using these scores, we modeled the potential for polygenic embryo prioritization within typical in vitro fertilization (IVF) scenarios. Our simulations indicate a relative risk reduction of 27-67% for the nine diseases for couples of European ancestry with five embryos. This demonstrates that polygenic prediction can translate into clinically relevant differences in prospective disease burden, even within the embryo numbers typically produced during IVF cycles.

Survey data of IVF patients demonstrates that there is interest in preimplantation testing for disease risk reduction. Indeed, the public imagination for this testing is far ahead of its actual use and, anecdotally, patients are often surprised to learn that disease risk reduction is not a routine element of embryo testing. For example, Furrer *et al*. surveyed 1427 individuals about their approval, interest, and concerns regarding various applications of embryo screening (25). Only 10.9% of those individuals disapproved or strongly disapproved of people using it, and only 11.3% believed that the risks of preimplantation genetic selection were greater than the benefits.

Clinical adoption has been cautious, in part due to the recommendations of scientific bodies like the American College of Medical Genetics and Genomics (ACMG). In 2024, the ACMG released a statement suggesting that the clinical utility of PGT-P to reduce disease burden remains unproven, and should not be offered as a clinical service (26). This was supported using a polygenic risk score for coronary artery disease (CAD) published six years before the statement was released, which explains only 41.8% of the variance of the CAD score developed within this paper. Obviously, weaker scores have poor clinical utility. The statement additionally indicates that the clinical utility of PRS testing may be affected by single-gene variants with large effects, and that a low PRS would be misleading in this instance. The ACMG suggests that including Preimplantation Genetic Testing for Monogenic disorders (PGT-M) would help solve this problem. The Nucleus processing pipeline addresses this concern by by including analysis of rare pathogenic and likely pathogenic variants across over 1800 genes, spanning all 84 genes recommended by the ACMG for reporting secondary findings in clinical exome and genome sequencing (27) and not reporting overlapping PRS results if such a variant is found.

Given the advances in PRS since 2018 and the addition of rare-variant analysis, we recommend the ACMG re-evaluate their position on PGT-P.

A frequently discussed benchmark in complex disease genetics is the proportion of heritability captured by a PRS relative to the theoretical maximum explainable by common genetic variation. This theoretical ceiling is typically represented by the common-SNP heritability, which quantifies the share of total phenotypic variance attributable to common variants tagged by SNP arrays.

Our three most predictive models–those for Alzheimer’s disease, prostate cancer, and type 2 diabetes–capture approximately 75% or more of the corresponding common-SNP heritability estimates (22,23,24). Thus, the scores cover a substantial proportion of the maximal common-variant signal currently detectable for these traits.

While these values remain below the broader heritability estimates derived from twin and family studies-which include effects from rare variants, gene-gene interactions, and shared environmental components–they nevertheless represent the upper bound of prediction achievable from common SNP data alone. Our aim is to eventually resolve the full heritability gap, while also optimizing predictive accuracy and clinical applicability within the limits of current genomic data.

Consistent with much of the published literature, our PRSs exhibit reduced predictive performance in individuals of non-European ancestry, particularly those of African descent. This attenuation reflects a well-recognized limitation of both current genomic resources and of the available polygenic risk methods (19). Although recent expansions of biobank-scale data from diverse populations–such as those from the All of Us Research Program and the Million Veteran Program–represent major progress, significant challenges remain. Future improvements will require large, ancestry-specific GWAS meta-analyses, more comprehensive LD reference panels, and modeling frameworks that explicitly integrate multi-ancestry summary statistics. Ongoing methodological developments, including trans-ethnic fine-mapping and ancestry-aware Bayesian models, may further close the gap in cross-population predictive accuracy.

Enhancing PRS performance for non-European populations is among our highest priorities. Achieving equitable predictive validity across ancestries is essential not only for scientific completeness but also for ensuring that clinical applications deliver benefits that are distributed fairly and globally.

The PRSs described here are immediately deployable within current clinical laboratory workflows for preimplantation genetic testing, provided appropriate counseling, regulatory oversight, and ethical safeguards are in place. As genomic reference data expand and diversity in GWAS improves, the predictive equity of these scores across ancestries will continue to strengthen, further increasing their utility in reproductive medicine.

## Methods

### Cohort Overview

We identified seven cohorts, five in AoU and two in UKBB. The five AoU cohorts included a European training cohort used for GWAS, and four non-European cohorts used for PRS testing. No individuals in the training cohort were related to individuals in the testing cohort above a King kinship coefficient of 0.0442, the lower bound of relatedness for third-degree relatives. The two UKBB cohorts included a European training cohort used for GWAS, and a European cohort used for PRS testing. No individuals in the training cohort were related to individuals in the testing cohort above a King kinship coefficient of 0.0442.

### AoU European Training Cohort

Within AoU, we defined individuals of majority European ancestry using AoU pre-computed majority genetic ancestry. Within our European cohort, we computed relatedness using King relatedness software implemented in Plink (12). From this, we identified a training cohort, which includes all individuals of majority European ancestry with electronic health records who have no relationship above a first-degree kinship coefficient of 0.177, or any relationship above a threshold of 0.0442 to any individuals with a first-degree relationship. The training cohort was used for GWAS in the development of our score. Next, we identified a subset of the training unrelated to each other above a King kinship coefficient of 0.0442, on which we ran 16 principal components (PCs) using the Plink –pca function. PCs were projected onto our training cohort for use in GWAS.

The whole-genome sequence cohort within the AoU Biobank were used for ancestry inference as well as GWAS. Variants which had missingness under 0.5%, minor allele counts greater than 100, and minor allele count to missingness ratios greater than 200:1 were retained, resulting in 7,205,124 variants overlapping with the reference panel.

### AoU Non-European Testing Cohorts

Within AoU, we identified individuals of majority African, Admixed American, East Asian, and South Asian ancestry using AoU pre-computed majority genetic ancestry. We combined these individuals with individuals of majority European ancestry, and computed relatedness using King relatedness software implemented in Plink (12). From this, we identified individuals within each non-European ancestry unrelated to our European training cohort or other individuals within their ancestry group above a King kinship coefficient of 0.0442, yielding four distinct test cohorts comprised of 52,023 individuals of majority African ancestry, 49,519 individuals of majority Admixed American ancestry, 6,410 individuals of majority East Asian ancestry, and 3,111 individuals of majority South Asian ancestry, respectively. We ran PCA generating 16 principal components (PCs) on each cohort using the Plink –pca function. PCs were projected onto our test cohorts for use in PRS validation.

### UK Biobank European Training and Testing Cohorts

Within the UK Biobank, we first defined a cohort of unrelated individuals using the pre-computed UK Biobank relatedness file. We ran 16 PCs over these samples using the Plink –pca function. We first denoted ancestry using self-reported data, establishing broad clusters of individuals who reported only European, African, Asian, Mixed, Other, and Unknown ancestry. We then projected PCs onto all whole-genome sequence (WGS) samples, computing PC centroids for individuals who reported only European, African, and Asian ancestry, and assigned individuals to the nearest centroid. We defined Europeans as individuals who were both genetically predicted to be European and reported only European ancestry,

We next used Plink implementation of KING relatedness software to compute relationships within our European cohort. From this, we identified three cohorts-first, a training cohort consisting of 40,872 siblings across 18,840 families. Second, we identified a training cohort of all individuals unrelated to our sibling cohort above a King kinship coefficient of 0.0442. Third, we identified a cohort of all individuals unrelated to both our sibling cohort and each other above a King kinship coefficient of 0.0442, on which we ran PCA generating 16 PCs using the Plink –pca function. PCs were projected onto our training and testing cohorts for use in GWAS and PRS validation, respectively.

The ML-corrected DRAGEN WGS dataset (Data-Field 24309) was used for ancestry inference as well as GWAS. Variants which were called as PASS, and had minor allele counts greater than 100, and missingness under 0.5% were retained, resulting in 7,319,488 variants overlapping with the SBayesRC variant panel.

### GWAS and Quality Control

The GWAS used in construction of our polygenic risk scores included internally-run UKBB and AoU GWAS, Finngen, MVP, and trait-specific consortia when available (28-31). GWAS participants were of majority European ancestry to match our LD panel. Internally-run AoU/UKBB GWAS were performed using Regenie, a machine-learning ridge based whole-genome regression with LOCO predictors to control for population stratification, on our population cohorts (32).

All summary statistics were harmonized to GRCh38 and filtered to remove variants with low imputation quality. For trait-specific consortium, variants suggesting low imputation quality or sample size were removed following recommendations from Tucker-Drob (33).

### PRS Construction

To construct our PRS, we first imputed each quality controlled summary statistics file using the SBayesRC impute function. The resultant summary statistics files were meta-analyzed together using inverse-variance weighting. The meta-analyzed imputed summary statistics were then used to construct PRS with SBayesRC, a bayesian hierarchical model which jointly models SNP effects in LD with annotation-informed priors. Resulting scoring files included weights from the 7,356,518 SNPs in the imputed summary statistics.

### PRS Liability R^2^ evaluation in UK Biobank

PRS validation must be performed in a target sample with no participant overlap or close relatives of any individuals used in the GWAS underlying the PRS. Sample overlap or close relatedness transmits associations between genetic information and environment, inflating performance metrics (34). PRS Liability R^2^ were evaluated within our predefined sibling cohort, which contains no samples related to our UKBB GWAS samples. From our sibling cohort, we randomly selected two individuals from each family and randomly assigned them to a cohort, creating two cohorts of 18,840 unrelated individuals. We computed liability R^2^ within each unrelated set using methods from Lee *et al*. (17) (35-43), and bootstrapped each set with 1000 replicates. We calculated combined R^2^ by meta-analysing each half with inverse-variance weighting.

### Within-Family PRS Evaluation within UK Biobank

To determine whether our PRS performance is attenuated within-family, we first fit a population model which regresses each binary phenotype on the standardized sibling PRS for that phenotype to estimate the population PRS coefficient *β_pop_*. We use a generalized linear mixed model with a binomial-probit link and a family random intercept, with age, sex, and the first 10 principal components as covariates, where *Y_if_* is the binary outcome for individual *i* in family *f*, Φ^−1^ is the probit link, α is the intercept on the latent scale, *X_if_* is that individual’s standardized PRS, C_*if*_ is the vector of covariates (age, sex, and principal components) and γ are their coefficients, and *u_f_* is a random family intercept.

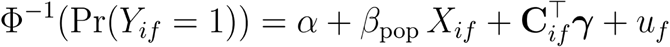

We next modified equation 1 from Selzam *et al*. (22), who decomposed PRS into a between-family mean *B_f_* and within-family deviation (*X_if_* – *B_f_*). We adapted this equation for binary phenotypes, fitting it with a probit generalized linear mixed model to estimate the within-family PRS coefficient *β_W_* while adjusting for the between-family component.

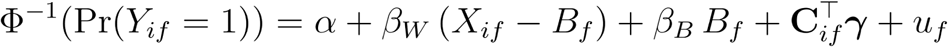

Models were fit in R using the lme4:glmer. We then computed the attenuation ratio *R_W:pop_* = *β_W_/β_pop_*. Standard errors were obtained via bootstrap with 1000 replicates.

### PRS evaluation in AoU Non-European Cohorts

PRS were evaluated within each of four non-European AoU v8 cohorts. We computed liability R^2^ using methods from Lee *et al*. (17). When ancestry-specific lifetime prevalences were unavailable, European lifetime prevalences were translated using odds-ratio or hazard-ratio scaling (40-47). Individuals who self-reported disease were classified as cases when used. Unless specified otherwise, individuals with 2+ disease code instances were considered cases, and individuals with 0 instances were considered controls, to more accurately distinguish cases and controls considering the varying medical coding accuracy across the United States (48).

### Calculation of Relative Risk Reductions

Relative risk reductions were calculated utilizing methods developed in Lencz *et al*. (5).

## Supplementary Tables

**Table 1.** GWAS Meta-Analysis Sample Sizes.

|  | Cases | Controls | Net-Effective Sample Size | Total Sample Size |
| --- | --- | --- | --- | --- |
| Alzheimer's Disease | 44,099 | 1,108,933 | 138,524 | 1,153,032 |
| Breast Cancer | 184,945 | 661,893 | 433,837 | 846,838 |
| Coronary Artery Disease | 243,541 | 1,256,267 | 774,954 | 1,499,808 |
| Endometriosis | 31,995 | 470,316 | 115,497 | 502,311 |
| Hypertension | 574,749 | 799,510 | 1,268,900 | 1,374,259 |
| Prostate Cancer | 161,032 | 824,745 | 432,759 | 985,777 |
| Rheumatoid Arthritis | 66,199 | 1,393,027 | 221,312 | 1,459,226 |
| Type 1 Diabetes | 7,020 | 977,850 | 27,824 | 984,870 |
| Type 2 Diabetes | 298,525 | 1,192,145 | 888,989 | 1,490,670 |

**Table 2.**
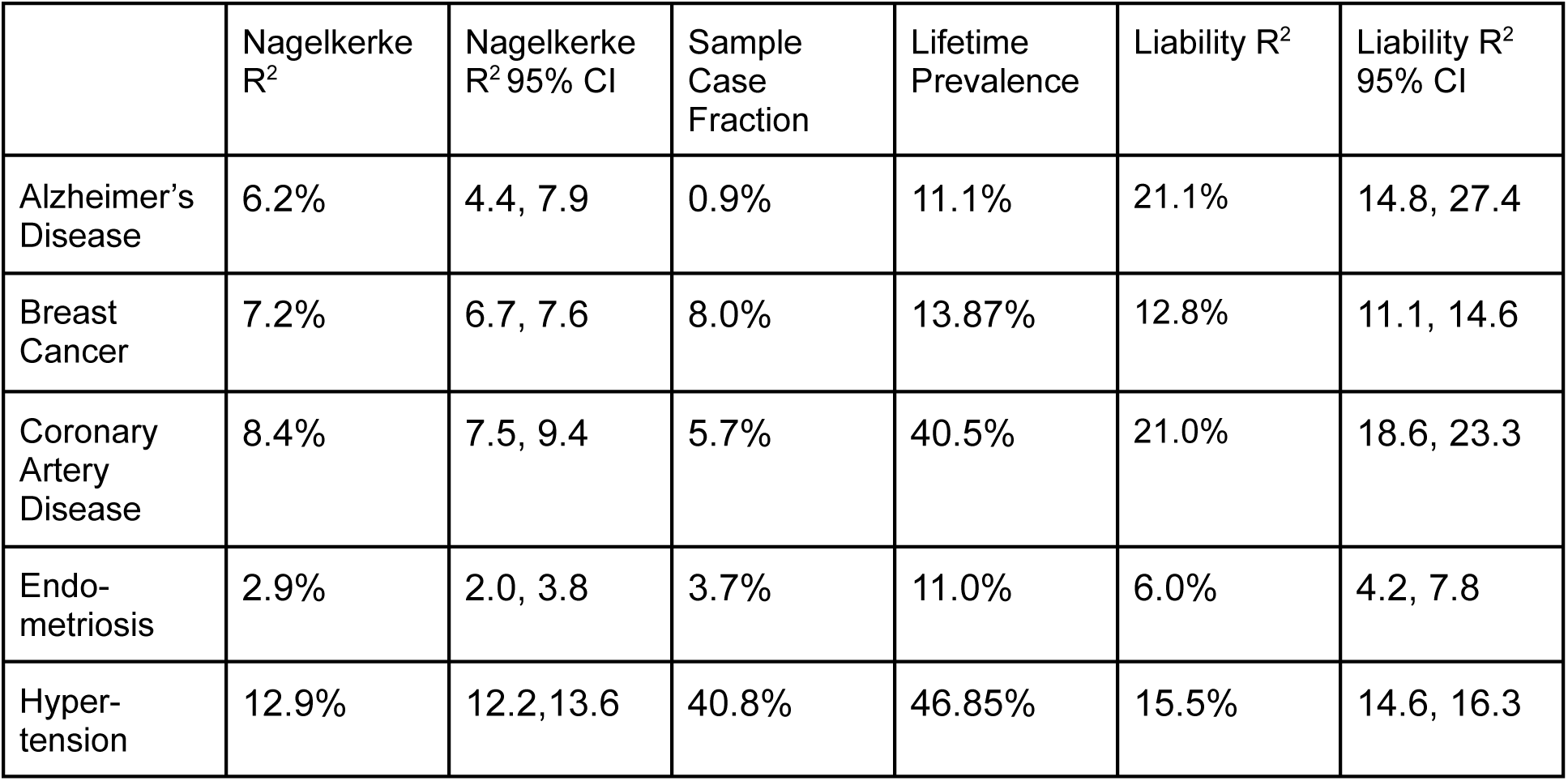
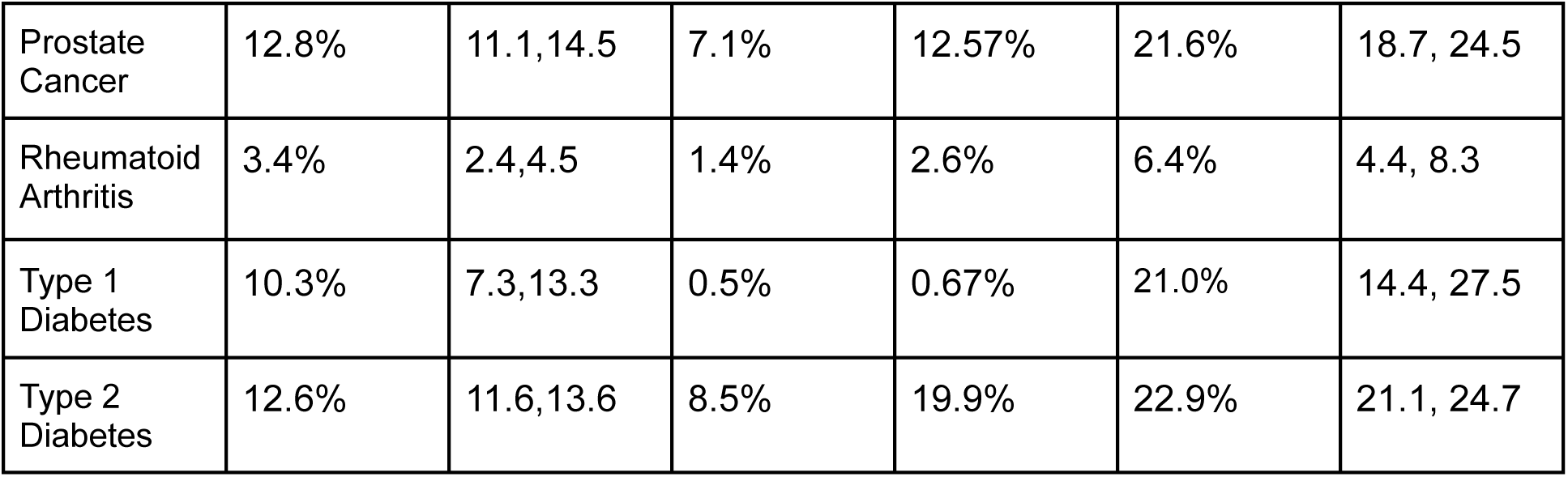
Nagelkerke and Liability-Scale R^2^ within UKBB.

**Table 3.** Within-Family Validation within UKBB.

|  | Within-family Beta | Population Beta | Beta Ratio | Beta Ratio 95% CI |
| --- | --- | --- | --- | --- |
| Alzheimer's Disease | 0.259 | 0.297 | 0.873 | 0.700, 1.045 |
| Breast Cancer | 0.331 | 0.363 | 0.911 | 0.804, 1.018 |
| Coronary Artery Disease | 0.399 | 0.393 | 1.014 | 0.944, 1.084 |
| Endometriosis | 0.209 | 0.204 | 1.026 | 0.781, 1.270 |
| Hypertension | 0.465 | 0.475 | 0.978 | 0.942, 1.014 |
| Prostate Cancer | 0.484 | 0.496 | 1.025 | 0.903, 1.146 |
| Rheumatoid Arthritis | 0.221 | 0.229 | 0.967 | 0.785, 1.148 |
| Type 1 Diabetes | 0.298 | 0.361 | 0.827 | 0.663, 0.991 |
| Type 2 Diabetes | 0.483 | 0.486 | 0.994 | 0.941, 1.048 |

**Table 4.** Phenotype Definitions.

|  | UKBB | AllofUs |
| --- | --- | --- |
| Alzheimer's Disease | 131036, 130836, 20002 (self-report code 1263) | ICD9: 331.0, ICD10: G30 |
| Breast Cancer | 40006 (ICD10 cancer code C50), 20001 (self-report code 1002) | ICD9: 174, 233.0, ICD10: C50, 836772 (self-report code 43528500) |
| Coronary Artery Disease | ICD9: 410-412, ICD10: I21, I22, I23, I24.1, I25.2<br>OPCS-4 codes (K40.1–40.4, K41.1–41.4, K45.1–45.5, K49.1–49.2, K49.8–49.9, K50.2, K75.1–75.4, K75.8–75.9), 20002 (self-report code 1075), 6150, 20004 (self-report codes 1070, 1095) | ICD9: 410-414, 429.7, 429.79, V45.81, V45.82, ICD10: Z95.1, Z95.5, I25.112, I25.83, I25.84 |
| Endometriosis | 132122, 20002 (self-report code 1402) | ICD9: 617, ICD10: N80 |
| Hypertension | 131286, 20002 (self-report codes 1065, 1072) | ICD9: 401-405, ICD10: I10, I11. Blood pressure at intake was evaluated as SBP+DBP+ 25 for hypertension medication presence. Total BP over 230 was considered as 1 case instance. |
| Prostate Cancer | 40006 (ICD10 cancer code C61), 20001 (self-report code 1044) | ICD9: 714, ICD10: C61, 836780 (self-report code 43529733) |
| Rheumatoid Arthritis | 131848, 20002 (self-report code 1291) | ICD9: 714.0, 714.1, 714.2, 714.81, ICD10: M05, M06 |
| Type 1 Diabetes | 2986 & (131848, 20002 (self-report code 1291)) | ICD9: 250.01, 250.03, 250.11, 250.13, 250.23, 250.31, 250.81, 250.83, 250.91, 250.93, ICD10: E10, O24.0-O24.03, & started insulin within 1 year of diagnosis & no Type 2 Diabetes Medication. |
| Type 2 Diabetes | 130708, 20002 (self-report code 1223) | ICD9: 250.00, 250.02, 250.20, 250.22, 250.30, 250.32, 250.40, 250.42, 250.50, 250.52, 250.60, 250.62, 250.70, 250.72, 250.80, 250.82, 250.90, 250.92, ICD10: E11, O24.1-O24.13. Individuals with 2+ code instances or 1 instance with 1 medication code were classified as cases, individuals with 0 code instances were classified as controls. |

## Acknowledgements

We gratefully acknowledge All of Us and UK Biobank participants for their contributions, without whom this research would not have been possible. Similarly we acknowledge all the participants in the cited GWAS articles that have contributed to the summary-level statistics used in this study.

## Datasets

This research has been conducted using the UK Biobank Resource under Application Number 331567, accessed using the DNAnexus cloud computing platform.

This research has been conducted using genetic and phenotypic data from the AllofUs Research Program’s Controlled Tier Dataset version 8, accessed using the AllofUs Researcher Workbench.

## Data availability

All PRS developed in this project are available through application at the Genetic Optimization Hub (https://mynucleus.com/labs/genetic-optimization-hub).

## Author Contributions

SC, KS, LF contributed to conceptualization; SC performed formal analysis; SC, PM, LF performed validation; LF, KS provided project supervision and data acquisition. All authors contributed to writing, review & editing and critical revision for the final manuscript.

